# Physical activity promotion is lacking in local treatment protocols for patients hospitalized with myocardial infarction: A cross-sectional study

**DOI:** 10.1101/2021.05.05.21256684

**Authors:** R. van Oorsouw, N. Koenders, J.P. Seeger, T.J. Hoogeboom

**Author notes:** Corresponding author R. van Oorsouw, PT, MSc^1^, Radboud university medical center, Radboud Institute for Health Sciences, Department of Rehabilitation, Nijmegen, the Netherlands.

## Abstract

**Objectives:** To describe the content and methodological quality of local physiotherapy and nursing treatment protocols specifically regarding physical activity promotion in patients hospitalised with myocardial infarction.

**Design:** Cross-sectional study comprising the analysis of 18 physiotherapy and 14 nursing protocols.

**Setting:** Protocols from twenty Dutch hospitals were used.

**Main outcome measures:** Data were extracted from protocols for patients hospitalised with myocardial infarction according to a standardized data-extraction procedure. Descriptive statistics were used to describe the content of protocols, in specific, treatment goals, interventions, clinimetrics and discharge criteria. Methodological quality of the protocols was examined using the AGREE-II tool.

**Results:** Therapeutic goals concerning physical activity were described in 83% of the physiotherapy protocols and in 7% of the nursing protocols. Therapeutic interventions concerning physical activity were described in 100% of the physiotherapy protocols and in 57% of the nursing protocols. Two (14%) of the fourteen interventions described in the physiotherapy protocols and none in the nursing protocols, were interpreted as physical activity promotion. All protocols were rated as having low methodological quality, mainly due to a lack of supporting scientific evidence. The overall quality was rated with an average score of 3 out of 7 (range: 1-5).

**Conclusions:** Physiotherapy and nursing protocols for hospitalized patients with myocardial infarction are primarily aimed at physical activity under supervision, rather than physical activity promotion. Our work provides physiotherapists and nurses insights in how to further improve the content and methodological quality of their local treatment protocols for patients hospitalised with myocardial infarction.

## Introduction

Physical inactivity is common among hospitalised patients, as they spend, on average, 51-83% of their hospital time lying in bed.[1–3] In the acute phase of illness or after surgery, it seems inevitable that physical activity levels drop. However, low physical activity levels are also found in patients who were able to be physically active.[2, 3] Unnecessary bed rest can delay recovery and harm patients.[4] Physical inactivity during hospital stay can lead to deconditioning with muscle weakness, reduced pulmonary function, and functional decline as important adverse effects.[5–7] The last couple of years increased awareness has been raised on the importance to end the epidemic of physical inactivity among hospitalized patients.[1, 8]

While being physically active is important for all hospitalised patients, it might especially be beneficial to patients hospitalised for MI. Evidence suggests that patients with myocardial infarction (MI) have an increased mortality risk when they delay ambulation.[9] International guidelines recommend professional support with early ambulation for patients hospitalised with MI, tailored to individual needs and preferences with help of the FITT criteria (Frequency, Intensity, Time and Type),[10–12] under the condition that the patient’s safety is ensured.[13]

Physical activity promotion contains all non-therapeutic actions to encourage and support increases in physical activity behaviour in individuals,[17] [18] and should ideally be utilized in all three phases of cardiac rehabilitation: hospitalisation and early rehabilitation (phase 1), outpatient rehabilitation (phase 2), and after care (phase 3).[19, 20] It the first phase, hospitalization and early rehabilitation, primarily physical therapists and nurses are the most logical candidates to promote physical activity. It is however, unclear to what extent physical activity promotion has been integrated in patient care of physical therapists and nurses, alongside their usual care.[21, 22] In the Netherlands, hospitals use local protocols to standardize care processes in line with international standards.[15, 16] These protocols describe, amongst others, goals, interventions, recommended clinimetric instruments, and discharge criteria. Information from the content and quality of local treatment protocols might provide insight into the current state of stimulating physical activity for people hospitalized with MI. Therefore, our aim is to describe the content and methodological quality of local physiotherapy and nursing treatment protocols specifically regarding physical activity promotion in patients hospitalized with myocardial infarction.

## Methods

### Design and eligibility

In this cross-sectional study, we asked department managers to share their local physiotherapy and nursing protocols for patients hospitalized with MI. Data were collected between September and November 2017. We used purposive sampling, targeting both academic and general hospitals, to gather protocols from all hospitals across twelve districts of the Netherlands. To maximise the response rate, we contacted hospitals through Dutch hospital physiotherapy networks: the association for hospital physiotherapy (NVZF) and the association for physiotherapy managers in intramural physiotherapy (VLF). Protocols were eligible for inclusion if they met the following criteria: 1) treatment protocols were developed for use by physiotherapists and/or nurses, and 2) for the treatment of patients hospitalised with MI. Reporting of this study followed the STROBE statement.[23]

### Data extraction

We used a standardised data extraction form to extract data on the content of the local, based on the work of Janssen et al.[24] First, all data regarding goals, interventions, clinimetric instruments, and discharge criteria were extracted. RvO and TJH, determined, independent of each other, which of the extracted goals and interventions were specifically related to physical activity promotion and which to regular mobilisation care. To our knowledge, there is currently no consensus on the definition of physical activity promotion, therefore, based on the work of both King and Stewart, we defined physical activity promotion as: all actions to encourage and facilitate physical activity in individuals or in the community, beyond the scope of an individual treatment session.[17, 18] Examples of physical activity promotion are motivational interviewing to increase physical activity behaviour or discussing a plan to achieve physical activity milestones after hospital discharge with shared decision-making.

### Methodological quality assessment

Methodological quality of the protocols was independently assessed by two trained physical therapists using the AGREE-II tool and checked by two authors (NK and TH).[25] This tool consists of 23 items that can be scored on a seven-point scale (1= ‘strongly disagree’ to 7= ‘strongly agree’), resulting in a total score between 23 and 161. There are six domains: 1) scope and purpose, 2) stakeholder involvement, 3) rigour and development, 4) clarity of presentation, 5) applicability, and 6) editorial independence. The overall quality was rated on a 7-point scale (‘lowest possible quality’ to ‘highest possible quality’). The AGREE-II tool has shown to be reliable and valid when used by physical therapists to assess clinical guidelines.[26] In case of discrepancies, the outcomes were discussed and if necessary, a third assessor (NK) was consulted.

### Statistical methods

Content of the protocols was presented using descriptive statistics. Data from physiotherapy and nursing protocols were compared using descriptive statistics. We calculated a quality score for each of the six domains of the AGREE-II tool.[25] The domain scores per question were calculated:

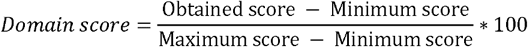

Furthermore, we calculated the average scores and standard deviations for all protocols and each domain score. A domain score of ≥60% was considered positive.[27, 28] If five or more domains of one treatment protocol scored >60%, the methodological quality was rated as good; if three or four domains of one treatment protocol scored >60%, the methodological quality was rated as neutral; and if less than three domains of one treatment protocol scored >60%, the methodological quality was rated low.[28]

## Results

In total, department managers of 61 hospitals were contacted of which 22 (36%) responded to our request. Two of the responding managers were not able to send the requested protocols, as one of them worked in a hospital without a cardiology ward and another one shared an outpatient protocol, which therefore was excluded. Ultimately, twenty different hospitals participated in this study; seventeen general and three academic hospitals. Since not all hospitals provided both their nursing and physiotherapy protocols, a total of eighteen physiotherapy protocols and fourteen nursing protocols were retrieved. Department managers of twelve hospitals sent both their physiotherapy and nursing protocol.

### Content of protocols

Goals concerning rehabilitation were described in 83% of the physiotherapy protocols and in 7% of the nursing protocols. Interventions concerning physical activity were described in 100% of the physiotherapy protocols and in 57% of the nursing protocols. Clinimetric instruments were described in 44% the physiotherapy protocols and in 21% of the nursing protocols. Discharge criteria were described in 50% of the physiotherapy protocols and in 50% of the nursing protocols. Hospital and protocol characteristics are displayed in Table 1. The differences in treatment goals, interventions, clinimetric instruments, and discharge criteria between physiotherapy and nursing protocols are displayed in Table 2.

**Table 1.** Hospital and protocol characteristics.

| Hospital | Hospital type | Physiotherapy protocols N = 18 |  |  |  |  |  | Nursing protocols N = 14 |  |  |  |  |  | Hospital covered all four domains |
| --- | --- | --- | --- | --- | --- | --- | --- | --- | --- | --- | --- | --- | --- | --- |
|  |  | Available | Created | Goals | Inter-ventions | Clinimetric instruments | Discharge criteria | Available | Created | Goals | Inter-ventions | Clinimetric instruments | Discharge criteria |  |
| 1* | General | A | - | + | + | - | + | A | - | - | - | - | - | - |
| 2* | General | A | 2017 | + | + | + | + | A | 2016 | - | - | - | - | + |
| 3* | Academic | A | - | + | + | + | + | A | 2011 | - | - | - | + | + |
| 4* | General | A | 2016 | + | + | - | - | A | - | + | + | - | - | - |
| 5* | General | A | - | + | + | - | - | A | - | - | + | + | + | + |
| 6* | General | A | - | + | + | - | + | A | - | - | + | + | + | + |
| 7* | General | A | - | - | + | - | - | A | 2015 | - | + | - | + | - |
| 8* | General | A | - | + | + | - | - | A | - | - | - | - | - | - |
| 9* | General | A | - | + | + | - | - | A | - | - | + | - | + | - |
| 10 | General | A | - | + | + | + | + | NA |  |  |  |  |  | + |
| 11 | Academic | A | 2017 | + | + | - | + | NA |  |  |  |  |  | - |
| 12 | General | A | 2017 | + | + | + | - | NA |  |  |  |  |  | - |
| 13 | General | NA |  |  |  |  |  | A | - | - | - | - | - | - |
| 14* | General | A | - | + | + | + | - | A | - | - | + | - | + | + |
| 15* | General | A | - | + | + | + | + | A | 2015 | - | + | - | - | + |
| 16 | General | A | - | - | + | + | - | NA |  |  |  |  |  | - |
| 17* | Academic | A | - | + | + | - | + | A | 2017 | - | + | - | + | - |
| 18 | General | A | - | + | + | - | - | NA |  |  |  |  |  | - |
| 19 | General | A | - | + | + | + | + | NA |  |  |  |  |  | + |
| 20 | General | NA |  |  |  |  |  | A | - | - | - | + | - | - |
| Totals | 17 General<br>3 Academic | 18 A<br>2 NA |  | 16<br>(83%) | 18<br>(100%) | 8<br>(44%) | 9<br>(50%) | 14 A<br>6 NA |  | 1<br>(7%) | 8<br>(57%) | 3<br>(21%) | 7<br>(50%) | 8<br>(57%) |
| Abbreviations: * Hospital sent both physiotherapy and nursing protocol A= available; NA = not available; + = described in protocol; - = not described in protocol |  |  |  |  |  |  |  |  |  |  |  |  |  |  |

**Table 2.**
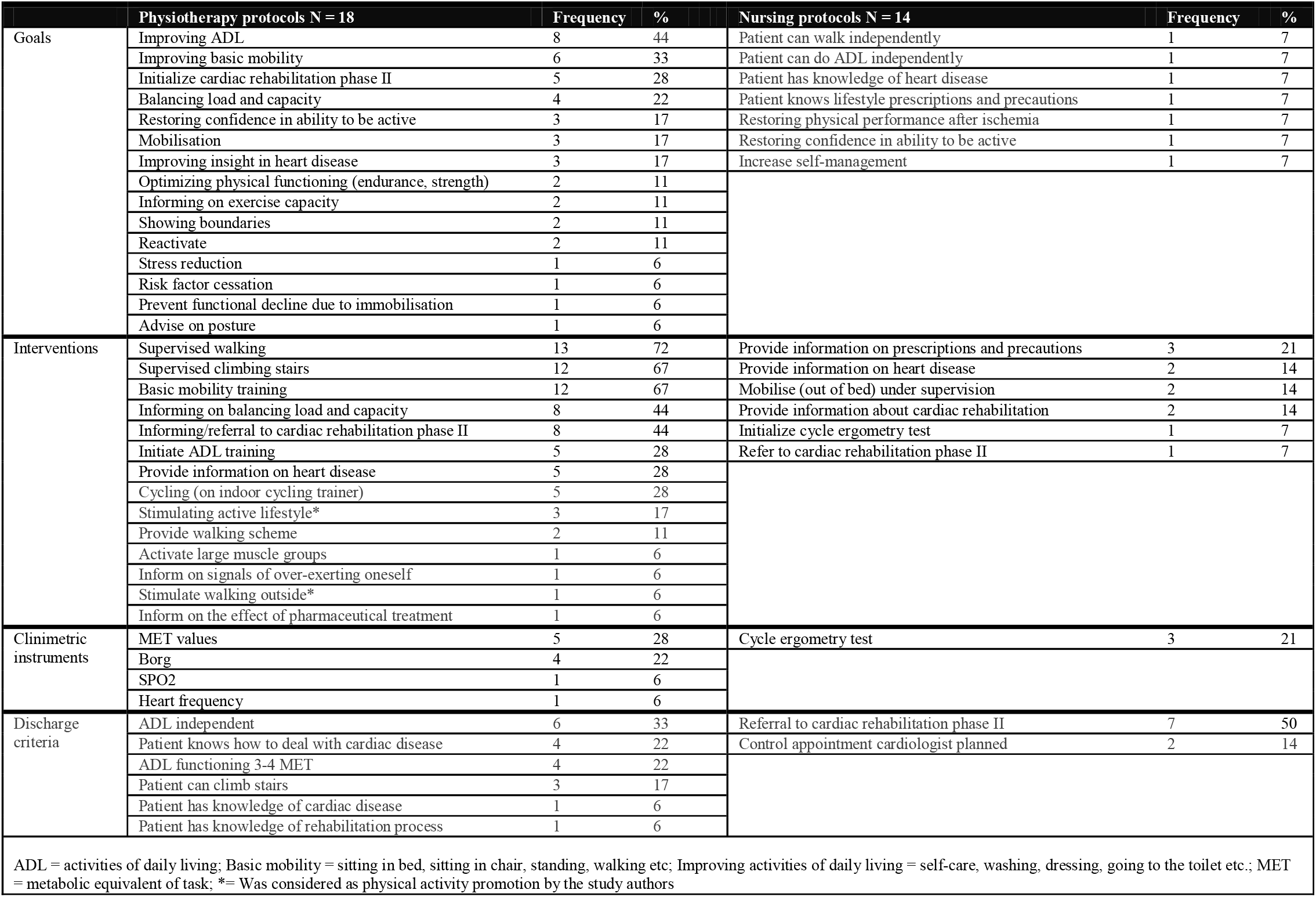
The content in treatment protocols concerning goals, interventions, clinimetric instruments and discharge criteria.

### Physical activity promotion in goals and interventions

None of the protocols described goals targeting physical activity promotion. Two (14%) of the fourteen interventions described in the physiotherapy protocols and none in the nursing protocols, were interpreted as physical activity promotion. Both these interventions aimed to stimulate physical activity: discuss active lifestyle and advise walking outside.

### Methodological quality assessed using the AGREE II - tool

The methodological quality scores of the treatment protocols are presented in Table 3. The first domain, Scope and Purpose, was scored positive in 33% of the physiotherapy (6 of 18) and in 36% of the nursing protocols (5 of 14). None of the physiotherapy or nursing protocols scored positive on any of the other domains (i.e., no local treatment protocol scored positive on two or more domains). Hence, all protocols were rated as low methodological quality. Domain scores per protocol are shown in Appendix 2. The overall methodological quality score of both physiotherapy and nursing protocols was rated, on average, 3 out of 7 (range: 1-5). (Appendix 2). The most frequently missing information were (from most to least frequent): Scientific evidence; Content of therapeutic actions; Goals and interventions; Measurements for physical activity; Discharge criteria; Poor clarity of presentation.

**Table 3:**
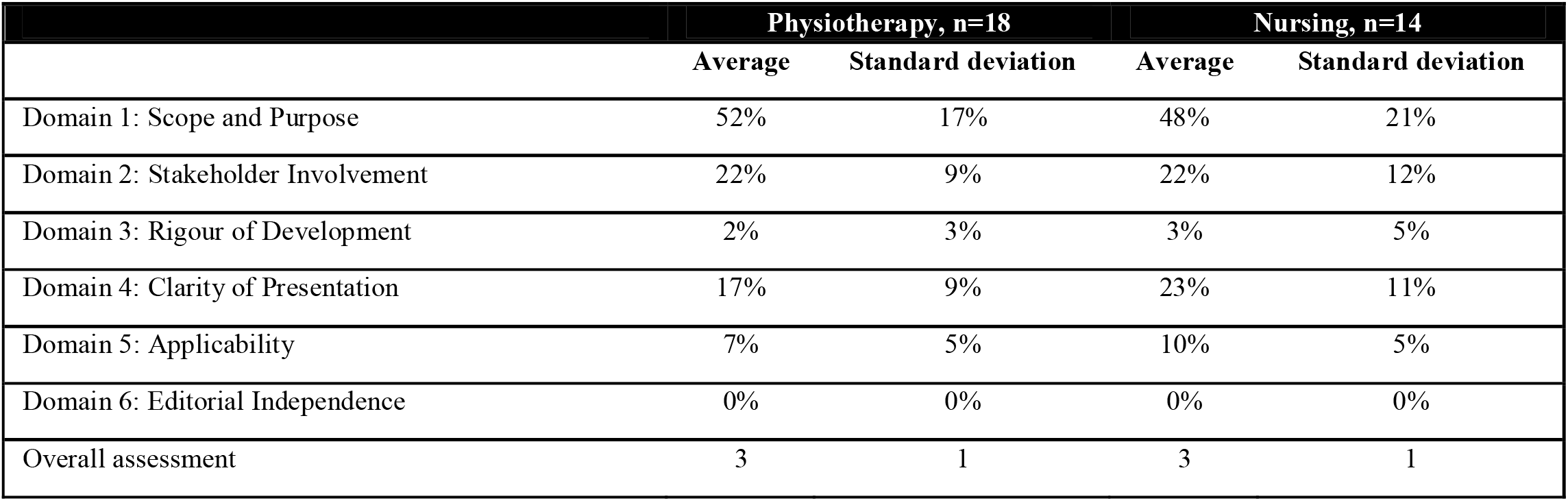
Outcomes of the methodological quality assessment with the AGREE-II tool for each domain and the overall assessment.

|  | Physiotherapy, n=18 |  | Nursing, n=14 |  |
| --- | --- | --- | --- | --- |
|  | Average | Standard deviation | Average | Standard deviation |
| Domain 1: Scope and Purpose | 52% | 17% | 48% | 21% |
| Domain 2: Stakeholder Involvement | 22% | 9% | 22% | 12% |
| Domain 3: Rigour of Development | 2% | 3% | 3% | 5% |
| Domain 4: Clarity of Presentation | 17% | 9% | 23% | 11% |
| Domain 5: Applicability | 7% | 5% | 10% | 5% |
| Domain 6: Editorial Independence | 0% | 0% | 0% | 0% |
| Overall assessment | 3 | 1 | 3 | 1 |

## Discussion

To our knowledge, this is the first study describing the content and methodological quality of local physiotherapy therapy and nursing protocols regarding physical activity promotion in patients hospitalised with MI. The majority of protocols missed essential content and clearly formulated treatment goals, interventions, and discharge criteria. Described goals and interventions aimed at rehabilitation or physical activity under supervision of healthcare providers, not at physical activity promotion. According to the AGREE II scores, methodological quality of all protocols included was considered low.

Nearly all physiotherapy protocols described goals and interventions addressing basic mobility such as sitting in bed, sitting in a chair, standing and walking. However, protocols frequently mentioned “mobilisation”, without providing detailed information on FITT criteria of interventions.[29] These criteria however are available and clearly described by the ACSM (Box 1). In line with international guidelines, the majority of the physiotherapy protocols described supervised walking, however the description lacked specificity. Clinimetric instruments were described in nearly half of the physiotherapy protocols, mostly including MET-values as advised in the Dutch guideline.[30] When clinimetric instruments were described they were not linked to treatment goals or discharge criteria. While for example the Borg-score could be used to measure exercise intensity and to provide instructions concerning physical activity.[31] Moreover, monitoring hearth frequency or hearth rhythm changes were only described in one physiotherapy protocol, while these vitals are needed to screen for contra-indications prescribing exercise, but also take an important part in applying FITT criteria.[11]

Nursing protocols contained hardly any information on physical activity nor its promotion. This is unexpected, as tasks such as stimulating and facilitating physical activity falls within the nursing domain, and the evidence about the nurses’ role in stimulating physical activity on hospitalised patients.[32] [33] The majority of the nursing protocols target restoring self-care in activities in daily living and providing medical information. These targets are only briefly named and not made explicit in goals or concise interventions. The importance of physical activity promotion for patients hospitalised with MI has not yet been translated in either the physiotherapy or nursing protocols. It raises the question: who claims the domain of physical activity promotion?

To our knowledge this is the first study assessing local hospital protocols using the AGREE-II tool, which was originally developed to assess clinical guidelines. We introduced this method because all items rated in the AGREE-II tool, despite the item “editorial independence”, are applicable to local hospital protocols. All protocols were scored as low methodological quality.[25] Domain 1 “Scope and Purpose”, showed the highest scores for both physiotherapy and nursing protocols. Objectives, health questions and population were described, but lacked specificity. The protocols scored low on all other domains, mainly because scientific evidence or substantiation of therapeutic actions was lacking and barriers and facilitators were not mentioned. This is in line with the findings of Oosting et al. (2009) who conducted a similar study with protocols for patients hospitalised with total knee replacement.[27] These authors also found poor methodological quality, lacking clinical reasoning and absent literature referencing. These findings indicate that physical activity is poorly described in hospital protocols and the information from applicable guidelines has not been integrated.

A limitation of this study is that we do not know to which extent protocols are implemented and used in hospitals. Therefore we cannot conclude that hospital care, concerning physical activity, is performed as described in the protocols. It might seem simplistic to assume that local protocols reflect the actual care as provided to patients. Moreover, these local protocols including all sorts of clinical management aspects and often focus on practical local arrangements. However, the role of protocols is to achieve good quality and responsible care for patients. (Inter)national quality systems require hospital professionals to follow protocols as these provide the staff and the organisation a standard to work with and to be audited by.[34] As local protocols are meant to standardise the care processes, this could mean that the quality of care is at stake. The delivery of either care might be lacking, certainly if protocols are used for longer spans of times and when newly appointed staff is trained (partly) with information from protocols. The quality of the treatment could be suboptimal, because protocols lack specific content despite availability of valuable literature on this topic. Further limitations of this study are the sample of protocols only from hospitals in the Netherlands, which means that the results might not easily be translated to the hospital context in other countries. However, the Dutch health care system is known as high quality and effective primary and secondary care,[35] which you would expect to be reflected in protocols. Moreover, the limited number of protocols in the sample enabled the authors to analyse the protocols in detail.

Findings from this study imply that there is a gap between information available in international guidelines and the content of local hospital protocols. Adopting this knowledge to local procedures could improve the implementation of physical activity promotion for hospitalised patients with MI. Furthermore, when physical activity promotion in a broader sense is specified in protocols it might raise awareness on the role of different healthcare providers.

## Conclusion

Physiotherapy and nursing protocols for hospitalized patients with myocardial infarction are primarily aimed at physical activity under supervision, rather than physical activity promotion. Information from international guidelines stressing the importance of physical activity promotion has not been integrated in these protocols. Our work provides physiotherapists and nurses insights in how to further improve the content and methodological quality of their local treatment protocols for patients hospitalised with MI.

## Data Availability

Data are available on reasonable request to the first author.

## Acknowledgements

We would like to thank Danique van Venrooij, Marijn Weren, Sabine van den Heuvel, and Femke Wolfs for their help during data collection and data-analysis. Furthermore we would like to thank the ‘Dutch association for hospital physiotherapy’ (NVZF) and the ‘association of managers in hospital physiotherapy (VLF) for their help contacting the hospitals. Finally, we want to thank the participating hospitals for sharing their protocols.

## Ethical approval

Not applicable.

## Box 1

**Frequency, Intensity, Time, and Type (FITT) criteria for early mobilisation after myocardial infarction.**

“**Frequency** = 2-4 sessions a day for the first 3 days of hospital stay; **Intensity** = Seated or standing HR_rest_ + 20 beats/min. Upper limit 120≤ beats/min that corresponds to a Borg-score ≤ 13 on a scale of 6-20; **Time** = Begin with intermittent walking bouts lasting 3-5 minutes as tolerated, progressively increase duration. The rest period may be a slower walk (or complete rest) that is shorter than the duration of the exercise bout. Attempt to achieve a 2:1 exercise/rest ratio. Progress to 10-15 minutes of continuous walking; **Type** = Walking, treadmill, or cycle. Resistance training is not recommended during the hospitalisation phase.”[11]

